# Comparing the accuracy of an ultrasound height measurement device with a wooden measurement board among children aged 2-5 years in rural Lao People’s Democratic Republic: a methods-comparison study

**DOI:** 10.1101/2023.07.21.23293003

**Authors:** Shan Huang, Caroline S.E. Homer, Joel Conkle, Sengchanh Kounnavong, Khampheng Phongluxa, Joshua P. Vogel

## Abstract

**Background:** Height is a key component of nutrition assessments in children from limited-resource settings. Traditional measurement boards are bulky and difficult to transport. We aimed to assess whether a handheld digital ultrasound device provides comparable accuracy to the measurement board for measuring children’s height.

**Methods:** We trained 12 health workers to measure the standing height of 222 children aged 2-5 years in rural Lao People’s Democratic Republic using the ultrasound device and measurement board. The Bland-Altman method was used to depict limits of agreement and potential bias. We reported the technical error of measurement (TEM) for precision, accuracy and assessed results against the Standardized Monitoring and Assessment for Relief and Transition (SMART) Manual 2.0 and the WHO Multicentre Growth Reference Study (MGRS).

**Results:** The average difference between the ultrasound and board measurements was 0.096 cm (95% limits-of-agreement: 0.041cm, 0.61cm) with a systematic bias of 0.1cm (95% confidence interval: 0.067,0.134), suggesting the ultrasound measurements measured slightly higher than those from the board. The ultrasound and board TEMs for precision were 0.157cm and 0.113 respectively. The accuracy TEM was 0.208cm. All TEMs were within SMART and WHO MGRS limits.

**Conclusion:** The ultrasound device is comparable to the measurement board among standing Lao children aged 2-5 years for precision and accuracy TEMs but showed a bias of 0.1cm. Further studies are required to assess whether calibration of device can minimise this bias and determine the ultrasound’s accuracy on recumbent length for infants and younger children.

## Introduction

Height and weight are basic anthropometric measurements that have long been used to create indicators of childhood nutrition status [1–3]. These two common anthropometric measures are used to monitor a child’s growth and development, as well as calculate subnational, national and international estimates of undernutrition and overnutrition. Estimation of the rates of stunting (low height-for-age) and wasting (low weight-for-height) in children under five years at a population level depends on accurate height and weight measurement [4]. These estimates are used by local and national governments to allocate resources for programs and activities to improve outcomes for childhood nutrition [5].

While digital scales are routinely used to measure a child’s weight, height measurements use measurement boards or stadiometers with readings done manually [6]. These boards are bulky, difficult to transport, costly, and prone to measurement errors in reading and recording reliable height measurements [7]. These measurement errors are attributable to their design and use, such as, incorrect positioning of the child against the board, difficulty in seeing measurements etched onto the board, the incorrect angle at which the measurement is read, and variability between different measurers [7]. Additionally, children aged under five years are challenging to measure due to their difficulty standing or lying still while measurements are taken [8].

Digital ultrasonic devices to measure length are used in the construction industry. Such devices are small, handheld, and potentially simpler to use compared to the bulky measurement board, however, their accuracy and precision in monitoring child growth have not been adequately reviewed or investigated. For ultrasound devices available on the market, we found no evidence regarding their use in children [9–11]. A few non-marketed devices have been formally tested in research settings with varying success [12–15]. Digital height measurement devices that are commercially available range from USD 25-50 per unit [9–11], making them significantly cheaper than measurement boards (at USD 114-259 each) [16]. If digital height tools were shown to be valid in clinical and survey settings, this could reduce costs of child height surveillance programs. Using global UNICEF procurement figures from 2012-2016, potential cost savings could be up to USD 3-7 million annually [7].

This study aimed to investigate the feasibility of a digital ultrasound device called One Grows™, to measure height in children aged two to five years old. We hypothesised that this device would be easier to use, clearer to read and show acceptable accuracy and precision in a limited-resource context when compared to the measurement board.

## Materials and Methods

### Study setting and participants

We conducted a method-comparison study in the district of Feung in Vientiane Province, Lao People’s Democratic Republic (Lao PDR). We aimed to recruit 220 children aged 2-5 years, who could stand independently without assistance. The intervention was to use the One Grows™ ultrasound device to measure the children’s height, and the control (comparison) was using a three-piece wooden height measurement board, the standard practice in Lao PDR. Both tools were used to measure children’s standing height only, not recumbent length. Participating children were identified and recruited from two kindergartens and three villages near the local health centre. We chose kindergartens and local villages as they are locations where health teams routinely perform community health outreach activities, including child height measurement in this age group [17]. Children were identified using convenience sampling, with written informed consent received from the child’s parent/caregiver prior to measurement.

### Study design and ethics

In 2017, UNICEF released a Target Product Profile (TPP) with recommendations for novel height measurement devices [7]. One criteria for new products was an accuracy of 0.3cm. We used this level of precision as the maximum allowable variation between the ultrasound and the measurement board. Applying this clinical delta of ±0.3cm, assuming a zero bias, 80% power, 95% confidence interval and 10% attrition/missing data, we calculated a sample size of 220 children. Using the Bland-Alman statistical methods for assessing agreement between two methods of measurement [18], this sample size allowed us to provide statistical inference for an approximate maximum standard deviation of the difference in measurements of 0.127cm and a 95% limit-of-agreement between the two methods of 0.249cm. This study was approved by the Ethics Review Board of the Alfred Hospital in Melbourne, Australia (ID: 142/22), and the National Ethics Committee for Health Research from the Ministry of Health, Vientiane, Lao PDR (ID: 2022.15). No identifying data from the participants were collected.

### Training on use of One Grows™ device and Standardization Exercise for measurement board

We trained 12 health centre staff from the Feung District and two study supervisors from the Lao Tropical and Public Health Institute (TPHI) on how to use the One Grows™ device. In Lao PDR, health centre staff have three years of tertiary level education and are trained as ‘Medical Assistants’. They are responsible for local public health activities, including maternal and child health promotion and community-based nutritional screening. As such, all those trained had previous experience using the measurement board but no experience with the ultrasound device.

The chief investigator (a public health nutritionist and clinical dietitian) delivered a two-day training. The training materials included a training manual on the application and maintenance of One Grows™ and a simple instruction card for easy reference during data collection. Aspects such as correct head and feet positioning (the same when using the measurement board) were key aspects of the training and well-practiced using the One Grows™ device.

Given previous experience, no additional training on using the measurement board was provided to the team. However, a standardisation test was administered to the 12 data collection team using the measurement board to assess how accurately and precisely measurers are able to use the board [1]. According to standardisation protocols, those who showed a Technical Error of Measurement (TEM) for both precision and accuracy under 0.6cm were considered to have passed this test [1]. Based on these results, six staff passed and became the measurers/enumerators for data collection. The remaining six staff became measurement recorders. In total, we had six measurement teams (one measurer and one recorder in each).

### Data collection

Data collection took place between 6-21 June 2022, immediately following the training. Written consent from the parent/caregiver was provided to the study team before any measurements were taken from the children. Children were selected in a random order to be measured. For each child, a total of six measurements were taken by a single enumerator – three measurements using the measurement board, and three using the One Grows™ device. The measurement process (Figure 2) alternated between the devices for each child to reduce the potential for recall bias should the measurer use the same device to measure the same child repeatedly one after another. The process then alternated in order again with the next child; for example, if child one was measured with the ultrasound device first, child two would be measured with the height board first and so on. Two study supervisors provided full time supervision and ensured consistency in following the measurement process for each child. They also monitored data quality and ensured accurate recording of measurements.

**Figure 1:**
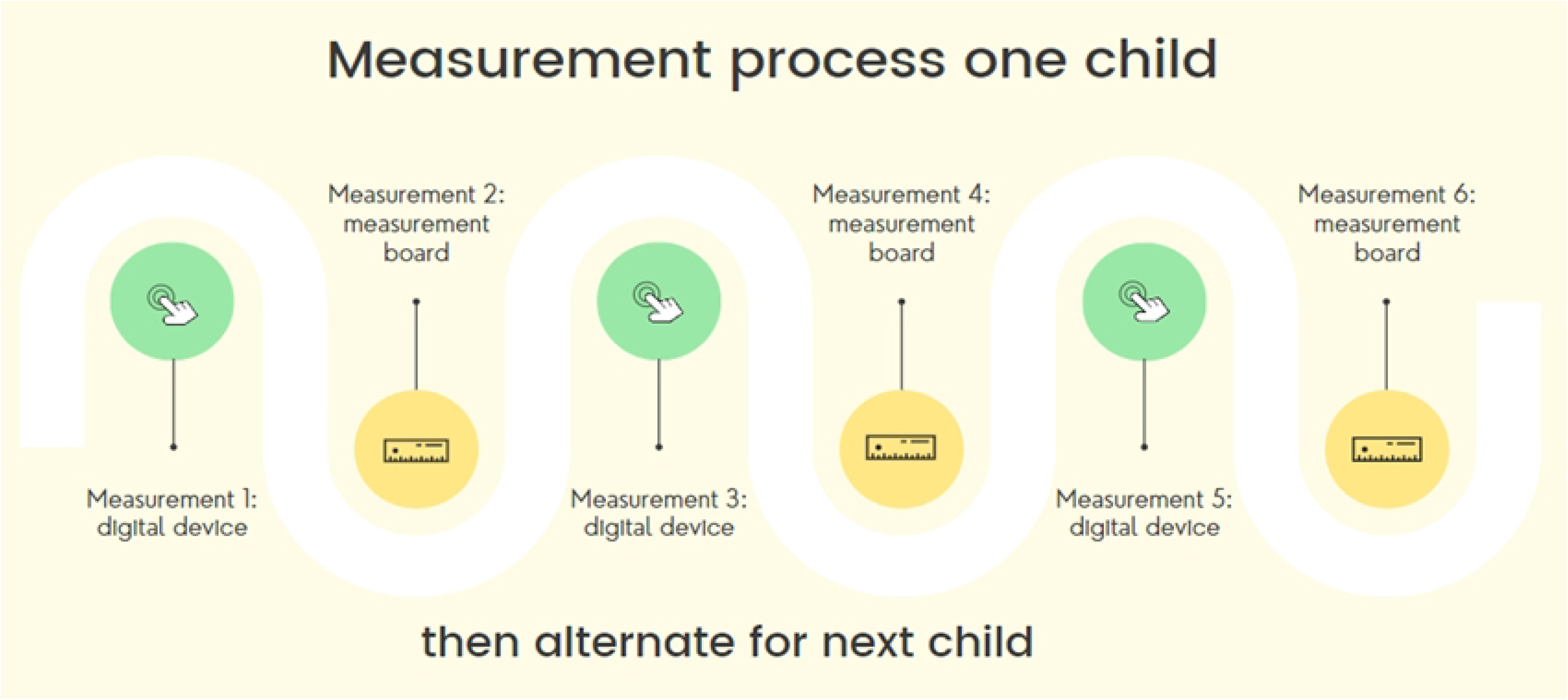
Measurement process for one child.

**Figure 2:**
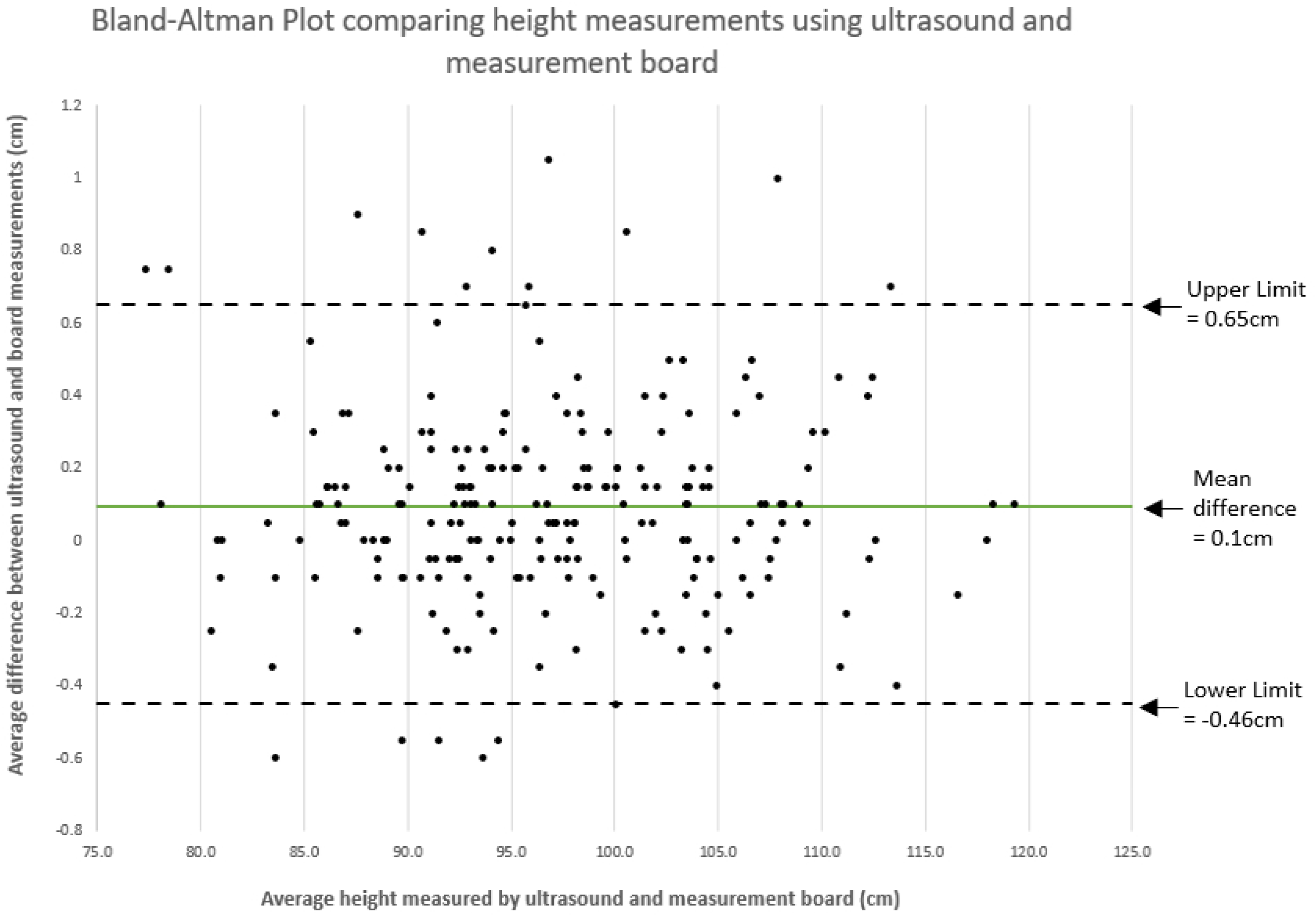
Bland-Altman Plot.

### Statistical methods

All measurements collected were recorded manually on a paper data collection form (one form per child), which were entered into a REDCap digital form by the study supervisors. All data were downloaded from REDCap and analysed using StataSE 17 software and Microsoft Excel.

We analysed our data based on the recommendations from the World Health Organization’s (WHO) Multicentre Growth Reference Study (MGRS) and the Standardized Monitoring and Assessment for Relief and Transition (SMART) Manual 2.0 [19, 20]. The MGRS created the global WHO Growth Standards, and provided guidance on how anthropometric measurements should be standardised. The data collectors and supervisors did not have these reference standards on hand during the data collection process. We followed the MGRS protocols for analysis where we took the first two measurements for both tools except where the first two measurements exceeded the maximum allowable difference of 0.7cm, the third measurement collected was used, and we analysed these measurements for precision and accuracy [19].

The precision and accuracy results were compared with the standards set by the SMART manual which represents the acceptable limits for the respective Technical Error of Measurements [20]. These are shown in Table 1. Both the MRGS and SMART are widely accepted to have set the standard in the way anthropometric measurements should be taken from children.

**Table 1:**
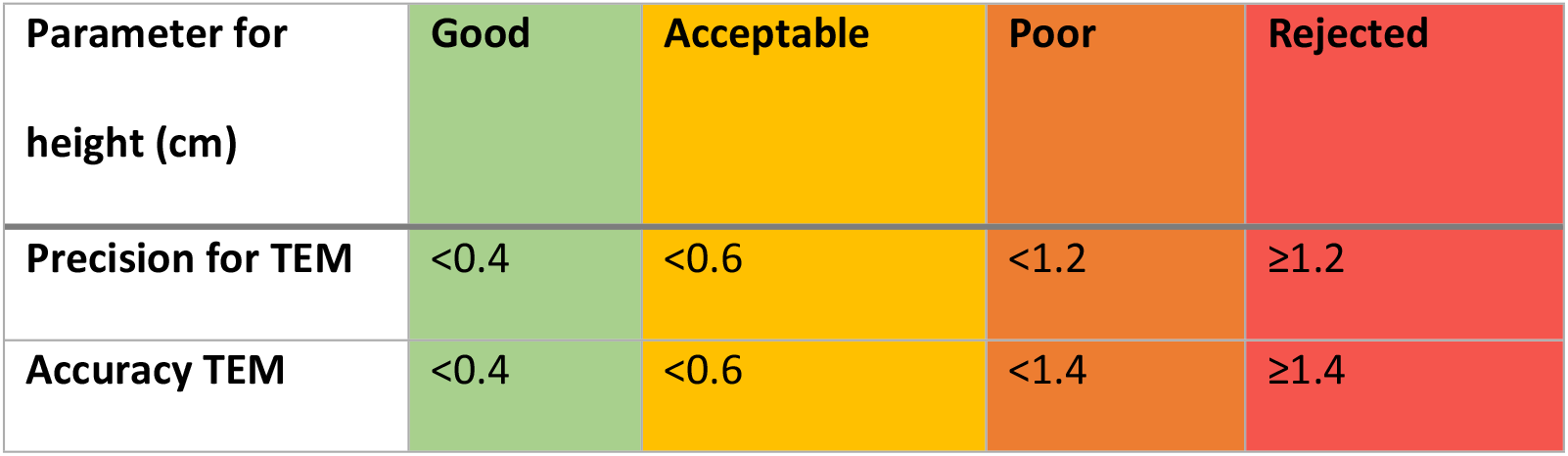
SMART Acceptable Limits for TEMs of precision and accuracy.

TEMs are commonly used in anthropometric assessments as a measure of accuracy and precision, and they can detect levels of variation from repeated measurements of the same individual, by the same measurer [21]. Its interpretation is that the differences between repeated measurements will be within ± the value of TEM two-thirds (66%) of the time, and 95% of the differences will be within ±2 ×TEM [21]. The lower the TEM, the smaller the variation in the repeated measurements. Hence, the TEM assesses the spread of measurements taken for the individual child being measured. The precision and accuracy were analysed by calculating TEMs (as per 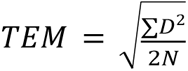 where D is the difference between the measurements taken and N is the total number of participants).

We define precision as the TEM between measurements taken with the same device to assess the consistency of the measurements taken. We define accuracy as the TEM between measurements taken with different devices, comparing the One Grows™ device to the measurement board.[20]

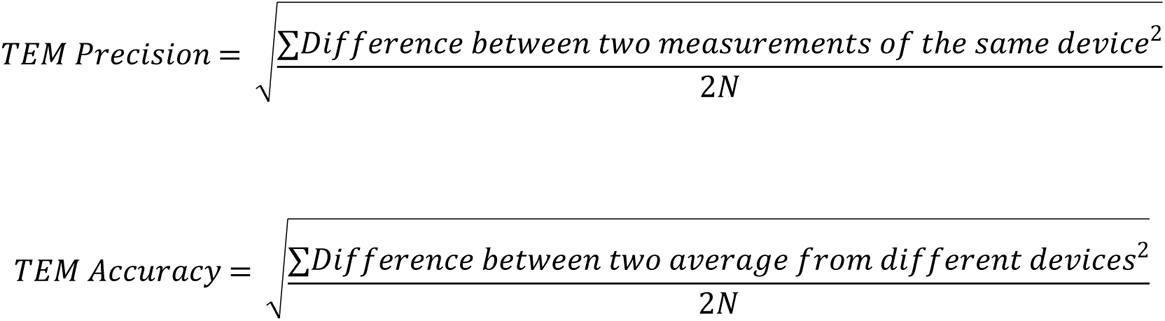

We used StateSE 17 for the Bland-Altman analysis and MS Excel to create the Bland-Altman plot. We compared the difference between the measurement devices across children of varying heights and ages, as well as the assessment of systematic bias. The Bland-Altman plot is a common way to represent results comparing two measurement methods [22]. The x-axis of the Bland-Altman plot is the average height measured by both the ultrasound device and the measurement board using the MGRS method, and the y-axis is the difference of the average ultrasound measurements subtracted from the average board measurements in centimetres. The mean difference between the average measurements for both devices was then calculated to assess systematic bias. That is, to assess how closely the ultrasound device is measuring to the measurement board. The upper and lower limits of agreement were plotted as two standard deviations from the mean difference of the two devices.

We used the Pitman’s test of difference in variance to determine if the difference between methods (y-axis) changed at different levels of average height (x-axis), where average height can be used as a proxy for age groups.

## Results

### Characteristics of study participants

In total, 222 children aged between 24-60 months (mean age 41.3 months) participated. Of the 222 children, 52.7% (n=117) were male and 47.3% (n=105) were female. For age, 20.3% of children were aged between 24-35 months (n=45), 39.6% aged between 36-47 months (n=88), 26.6% aged between 48-59 months (n= 59) and 13.5% aged at 60 months (n=30) (Table 2).

### Comparison of measurement methods using the Bland Alman method

The summary statistics are presented in Tables 3 and 4. The average of all three measurements taken using the digital ultrasound device was 97.04cm (range, 77.63 to 119.33cm; SD = 8.22cm) compared to the average of all three measurements taken with the measurement board of 96.94cm (range, 77.03 to 119.27cm; SD = 8.21cm).

**Table 3:** Summary statistics for all measurements taken using both devices (n=222 children).

|  | Mean | Standard<br>Deviation | Min<br>Measurement | Max<br>Measurement | Standard<br>Error |
| --- | --- | --- | --- | --- | --- |
| **Ultrasound measurements** |  |  |  |  |  |
| **Ultrasound Measurement 1 (cm)** | 97.06 | 8.22 | 77.90 | 119.30 | 0.55 |
| **Ultrasound Measurement 2 (cm)** | 97.04 | 8.21 | 77.60 | 119.40 | 0.55 |
| **Ultrasound Measurement 3 (cm)** | 97.03 | 8.23 | 77.40 | 119.30 | 0.55 |
| **Average of all Ultrasound**<br>**Measurements (cm)** | 97.04 | 8.22 | 77.63 | 119.33 | 0.55 |
| **Board measurements** |  |  |  |  |  |
| **Board Measurement 1 (cm)** | 96.96 | 8.21 | 77.00 | 119.20 | 0.55 |
| **Board Measurement 2 (cm)** | 96.93 | 8.22 | 77.00 | 119.30 | 0.55 |
| **Board Measurement 3 (cm)** | 96.93 | 8.21 | 77.10 | 119.30 | 0.55 |
| **Average of all Board Measurements (cm)** | 96.94 | 8.21 | 77.03 | 119.27 | 0.55 |

**Table 4:** Difference of measurements between ultrasound and board measurements (cm) using the MGRS method.

|  | **Mean difference** | **Difference range** | **Standard Deviation of difference** | **Limits-of-agreement** |
| --- | --- | --- | --- | --- |
| **Difference of measurements** | 0.1 | -0.60 and 1.05 | 0.28 | -0.46, 0.65 |

The Bland-Altman plot in Figure 2 indicates that the 95% limits-of-agreement between the board and the ultrasound device using the MRGS method was between −0.46cm to 0.65cm (SD = 0.28cm). There was a range of difference when we subtracted the ultrasound measurements from the board measurements, between −0.6 and 1.05cm. The Bland-Altman analysis using StataSE 17 calculated the overall mean difference, or systematic bias, to be 0.1cm (95%CI 0.06 to 0.13cm). This indicates that the ultrasound device was consistently measuring the children 0.1cm taller than the board in this population.

Pitman’s test results show that the difference between the board and the ultrasound device did not significantly differ by average height (r = 0.002, p = 0.973). This indicates that a child’s age had no relationship with the difference in height measurements between the two devices.

### Precision and Accuracy

#### Precision

As shown in Table 5, the precision TEM for the ultrasound device was 0.157cm (females: 0.157cm, males: 0.158m), and the precision TEM for the height board was 0.113cm (females: 0.101cm, males: 0.123cm). Based on the standards set by the SMART manual, TEMs for both ultrasound device and height board were <0.4cm, indicating ‘good’ precision.

**Table 5:** Precision and bias of ultrasound and measurement board.

|  | Females | Males | All |
| --- | --- | --- | --- |
| **Precision TEM Digital Ultrasound** | 0.157cm<br>(n=210) | 0.158cm<br>(n=134) | 0.157cm<br>(n=444) |
| **Precision TEM Measurement Board** | 0.101cm<br>(n=210) | 0.123cm<br>(n=134) | 0.113cm<br>(n=444) |
| **Accuracy TEM between Digital and Measurement Board** | 0.227cm<br>(n=210) | 0.190cm<br>(n=134) | 0.208cm<br>(n=444) |

#### Accuracy

The first accuracy calculation made using the two closest measurements between the ultrasound and the height board was 0.208cm (females: 0.227cm, males: 0.190cm). Similarly, based on the SMART standards, these bias levels also indicated ‘good’ accuracy between the two devices, with girls showing a slightly greater bias than boys.

## Discussion

This study of 222 children in Lao PDR compared measurements taken using the One Grows™ ultrasound device and the UNICEF wooden measurement board (standard of care). For reliability, the TEM results were within acceptable SMART and WHO MGRS limits and indicated that each device was consistent with itself. The measurement board showed a slightly higher precision than the ultrasound device (TEM 0.113cm vs 0.157cm).

For accuracy, the TEM of 0.208cm was well within the acceptable limits of the SMART and WHO MGRS guidelines [19]. There was a small difference in bias TEM among girls and boys - the bias TEM among girls was higher by 0.037cm. Upon closer inspection of the measurements recorded, among the ultrasound measurements, there were three occasions where the third measurement had to be used because the maximum allowable difference was exceeded among the girls, but only one occasion of this occurred when measuring the boys. None of the board measurements required adjusting (using a third measurement). During the study, we observed that girls often had long hair tied up in elaborate styles. It is possible that when the measurer releases a child’s hair to perform measurement, their hair is still messy and might cause additional movement (and hence additional differences between measurements) when using the handheld ultrasound device. Conversely, the headpiece of the measurement board is heavy and thus may be steadier. Further investigation of possible sources of measurement-to-measurement variation when using a handheld ultrasound device is required.

The ultrasound device produced a systematic bias of 0.1cm and was shown to measure higher than the measurement board. It is difficult to conclude that the ultrasound device is truly measuring children taller than their true height because the measurement board itself is not a gold standard, rather an operational standard since no other tools are currently used to measure height. Therefore, without proper calibration of either the ultrasound device or the board, it is not possible to say for certain whether the ultrasound device is measuring the child taller, or the board is measuring the child shorter. Future studies need to consider calibration of both devices.

When child height is measured in a clinical setting, usually only one measurement is taken. Although the board showed slightly higher precision, our findings suggest the ultrasound device can be used in clinical settings. Given the greater portability, lightweight nature, lower cost, and ease-of-use of the ultrasound device, it could be a better choice for clinical use, particularly in limited-resource settings.

The 2017 UNICEF TPP [7] suggested a novel height measurement device needed to meet two essential requirements: firstly, to improve upon currently available measurement boards with a digital output; and secondly to use innovative technologies (such as ultrasound, infrared or laser). The One Grows™ device meets both UNICEF TPP essential requirements, as well as several optional requirements including an associated mobile application (app) whereby the measurement taken by the device is automatically entered via Bluetooth technology. The app also allows for multiple measurements on multiple children. Our device also met the accuracy range set by the TPP, which was within 0.3cm for TEM. Our study showed that the One Grows™ device meets the main requirements from the UNICEF TPP except for a low battery indicator and assessments for commercialization requirements.

To our knowledge, this is the first study to compare this handheld ultrasound device with the measurement board. Few other ultrasound devices have been studied in detail on children, and none of these studies were done in the context of rural health workers in a low or middle-income country. In 1998, Watt, Pickering and Wales published a study with 18 children (ages unspecified) in the United Kingdom using the Gulliver G-100 ultrasound device, which showed an average bias between 0.74-0.88cm when compared with a Harpenden stadiometer [12]. In 1999, Glock et al also used the G-100 device to assess growth hormone treatment of 101 children with severe dwarfism in Germany, with results showing an average bias of 0.49cm when compared to the Harpenden stadiometer [14]. For both these studies, the difference in measurement between the devices were more than five times greater than our results (0.1cm vs. >0.49cm). In 2013, Syafiq and Fikawati [15] tested the feasibility of a prototype measurement board using an ultrasound attachment (the P2B2D) on 53 infants in Malaysia compared to a plastic length board. Their inter-method TEM of 3.66cm is rejected based on SMART standards [20]. It is noteworthy that Syafiq and Fikawati measured the length of infants, whereas our study measured standing height in children 2-5 years. Length is known to be more difficult to measure correctly than standing height, as it requires careful positioning to ensure the child is appropriately stretched before taking the measurement [3]. In 2020, Cho et al validated a handheld ultrasound device (InLab S50) among 100 adults in South Korea, this device is similar to the One Grows™ device used in this study, and reported similar findings.[13] When compared to the stadiometer, the InLab S50 had a mean bias of −0.15cm (95% limits-of-agreement: −1.69cm, 1.38cm), whereas in our study, the mean bias was 0.1cm (95% limits-of-agreement: −0.46cm, 0.66cm). By comparison, the InLab S50 consistently measured lower height compared to the board, while the One Grows™ measured greater than the board. Our study also showed smaller margins of difference compared to the InLab S50 device.

### Strengths and limitations

Our study demonstrated the One Grows™ ultrasound device performs valid height measurements in standing children 2-5 years of age. These measurements were accurate and as precise as wooden measurement boards, which has not been shown in the few previous studies evaluating ultrasound-based height measurement devices. Strengths of this study included a large sample size (222 children), adherence to the WHO MGRS methodology, and comparing our results to the SMART standards. An additional strength is novelty – this study is the first to evaluate an ultrasound device for child height measurement in a limited-resource setting, performed by healthcare personnel with limited formal training who routinely perform child nutritional surveillance activities. The study setting and training approach closely reflects the real-world needs for novel, digital devices. Hence, this study is a critical step in the investigation into how handheld height measurement devices can be used to measure child stature in limited-resource contexts. These results demonstrate that this device may be appropriate in place of the measurement board, though further research in other contexts is warranted.

While our study findings demonstrate that the two devices were comparable to each other, a limitation was that we only measured children who could stand at 2 to 5 years of age – we did not test it on children under two years where recumbent length is measured. We also note that the comparison TEMs are set against children aged 0-5 years, not children aged 2-5 years. For recumbent length, this ultrasound device would measure length from foot to head, rather than height from head to ground. This would require further investigation to determine if the accuracy is similar in recumbent measurement. Our measurement process of alternating device for the same child could contribute to measurers being biased by previous measurements. To reduce this bias, in future studies, it may be worth considering the measurement of an entire group of children once with both devices, then re-measuring the same group of children another time over. Our study design also did not permit assessment of intra-measurer reliability, as only one measurer performed all measurements for a single child. While we did not formally assess the cost-effectiveness of the ultrasound device, it is cheaper than the measurement board (USD 50 compared to USD 114-259 from the UNICEF Supply Catalogue) [7]. We consider that a formal cost-effective evaluation that considers durability and recurrent costs would be useful to guide procurement decision-making.

## Conclusion

In a methods-comparison study of 222 children between 2-5 years old in Lao PDR, the One Grows™ ultrasound device showed good levels of precision and accuracy in measuring height of standing children. However, there was a systematic bias between the devices, with the ultrasound measurement being 0.1cm greater than that of the measurement board. Future studies showing calibration of both devices will be required to ascertain which of these devices measures the closest to the true height/length of a child; and whether the ultrasound device will require further recalibration. The measurement board is bulky and heavy to carry, whereas this ultrasound device is handheld, lightweight, and cheaper. Additional studies involving measurement of recumbent height in younger children are required, as well as further assessments to explore intra-measurer reliability, and device performance in other settings.

## Data Availability

Data is presented in this study are opening available in Monash University Bridges (Figshare) online repository at DOI: 10.26180/22678663.

## Acknowledgements

The study would like to thank the Feung District Hospital and the Tropical and Public Health Institute in Lao People’s Democratic Republic for their data collection efforts and field work supervision. The study team would like to acknowledge the children, teachers and communities of the Son Samai, Chang Noi kindergartens and Phonexay, Nakang and Phonsavath villages for their involvement. The authors also acknowledges funding from the Burnet Institute to support this study.

## Supporting Information

**S1 Fig. Depiction of the measurement process**

**S2 Fig. Bland-Altman Plot**

